# Association between telecommuting environment and low back pain among Japanese telecommuting workers: A cross-sectional study

**DOI:** 10.1101/2021.07.15.21260610

**Authors:** Ryutaro Matsugaki, Keiji Muramatsu, Seiichiro Tateishi, Tomohisa Nagata, Mayumi Tsuji, Ayako Hino, Kazunori Ikegami, Yoshihisa Fujino, Shinya Matsuda, for the CORoNaWork Project

## Abstract

**Objectives:** We evaluated the relationship between telecommuting environment and low back pain (LBP) among desk-based workers in Japan.

**Methods:** This cross-sectional study included 3,663 desk-based, telecommuting workers. LBP was assessed using a 0–10 numerical rating scale. The telecommuting environment was evaluated using subjective questions. Mixed-effects logistic regression analysis was used to evaluate this association.

**Results:** The results of mixed-effects logistic model revealed that not having a place or room to concentrate on work, desk not well-lit enough for work, lack of space on the desk to work, not having enough legroom, and uncomfortable temperature and humidity conditions in the workspace were significantly associated with higher odds of LBP.

**Conclusions:** Our findings suggest that telecommuting environment is associated with the prevalence of LBP.

## Introduction

The coronavirus disease (COVID-19) pandemic has led to a rapid expansion of telecommuting in Japan. Moreover, the Japanese government has recommended telecommuting to prevent the spread of COVID-19^1,2^. Although the telecommuting rate among Japanese workers was 14.8% as of October 2019, it increased to 23.0% by November 2020^3^. In actuality, 85.0% of workers who were teleworking as of November 2020 did so to prevent the spread of COVID-19^3^. Telecommuting is a way to continue business even during times of emergencies, and it may even become a more common way of working.

With the rapid expansion of telecommuting due to the COVID-19 pandemic, management of the work environment while telecommuting is an emerging occupational health problem. As with the office environment, the recommended working environment for a telecommuting worker is a private, quiet, and safe dedicated space, with adequate lighting, comfortable temperature and humidity, ergonomic chairs/desk^4–6^. However, it is difficult to manage the telecommuting work environment because, in contrast to the office, the workstation at home is difficult to supervise by managers, and managers lack the authority to direct the private environment of an individual. In addition, neither companies nor workers were sufficiently prepared to optimize a telecommuting environment due to the unexpected occurrence of COVID-19^4^. It has been reported that more than 50% of telecommuting workers in Japan lack a desk/chair and more than 70% lack a private room or space for work^3^.

Low back pain (LBP) is an important health problem associated with office work. The prevalence of LBP is 34-56% among office workers^7–10^. In office workers, gender, body mass index, sleep disturbance, and previous symptoms of LBP are known to be risk factors for LBP^8,10–13^. In terms of work-related factors, sitting time and sitting posture at work are known risk factors for LBP among office workers^8,14,15^. At home, the work environment is less developed than that in the office environment; therefore, the work environment of home workers is assumed to be an important risk factor for LBP. Working in a space that is unsuitable for work can induce the worker to take awkward postures, which may have a negative impact on the musculoskeletal system.

Previous studies conducted during the COVID-19 pandemic have suggested an association between telecommuting and musculoskeletal pain^16^. However, the relationship between work environment and LBP in telecommuting workers remains unclear. Thus, the purpose of this study was to clarify the relationship between the work environment and LBP in telecommuting workers.

## Methods

### Study Design and Subjects

This cross-sectional Internet-based monitoring survey was conducted from December 22 to December 26, 2020, when the third wave of COVID-19 began in Japan. The details of the survey protocol have been previously reported^17^. Data were collected from workers with employment contracts at the time of the survey. Among 33,302 workers, 27,036 were surveyed, excluding those who gave fraudulent answers. Among those surveyed, 3,663 (2,093 males and 1,570 females) workers, who responded that they mainly performed desk work (e.g., office work, computer work) and telecommuted at least once a week, were included in the present analysis.

This study conformed to the principles of the Declaration of Helsinki. In addition, the study was approved by the ethics committee of the University of Occupational and Environmental Health, Japan (reference no. R2-079). Informed consent was obtained online from the participants through the website.

### Assessment of LBP

We assessed the presence of LBP in the participants based on two questions. First, we asked all subjects “Have you experienced stiff shoulders or LBP in the past two weeks?” and asked them to answer, “yes” or “no.” If the subject answered “yes” to that question, then the following questions were asked to assess the severity of LBP, such as “what was your average level of LBP in the past 2 weeks? (Please rate your pain from 0 to 10, where 0=no pain at all and 10=the most intense pain you have experienced).” The numerical rating scale (NRS) was used for evaluating pain severity. In this study, a score of 3 or higher on the NRS indicated the presence of LBP, similar to previous studies^18,19^.

### Assessment of the telecommuting environment

The telecommuting environment was assessed using the following questions: 1) “Is your desk well-lit enough for you to work?”; 2) “Is there enough space to stretch the legs?”; 3) “Do you have a place or room where you can concentrate on your work?”; 4) “Are the degrees of temperature and humidity in the room where you work appropriate for working comfortably?”; 5) “Do you have enough space on your desk to work?”; 6) “Do you use an office desk or chair (including children’s study desks)?” The respondents answered either “yes” or “no.” These items were designed according to the guidelines for the promotion of appropriate introduction and implementation of telework by the Ministry of Health, Labour and Welfare^6^.

### Assessment of participants’ characteristics and other covariates

The following socioeconomic factors were examined: age, sex, body mass index (calculated by dividing the weight by height squared), educational background (junior high school, high school, vocational school, junior college or technical college, university, or graduate school), and equivalent income (household income divided by the square root of the household size).

The following lifestyle factors were examined: smoking (currently smoking), drinking (alcohol consumption on two or more days per week), physical activity (perform equivalent physical activities for at least 1 h a day in daily life for more than 2 days a week), and exercise habit (exercise for 30 minutes or more for more than two days a week).

Mental health status was assessed using the following question: “During the past 30 days, for how many days did you experience poor mental health, including stress, depression, emotional problems, etc.?”. This item is part of the Centers for Disease Control and Prevention core healthy day measures assessing health-related quality of life (CDC HRQOL-4)^20^. The Japanese version of CDC HRQOL-4 has been validated^21^.

The following work-related factors were examined: work duration (hours per week), frequency of telecommuting (one day per week, more than two days per week, and more than four days per week), and company size (total number of employees at the company where the respondent works).

### Statistical analysis

Age, body mass index, and number of days the participant experienced poor mental health during the past 30 days were expressed as continuous variables, and data are presented as mean and standard deviation. Other variables were expressed as categorical variables and presented as numbers and percentages.

Mixed-effects logistic regression analysis was conducted with LBP as the dependent variable, subjective evaluation of the telecommuting environment as the independent variable, and the city of residence as the random effects. We used age, sex, body mass index, lifestyle habits, number of days of poor mental health, equivalent income, educational background, working time, frequency of telecommuting, and company size as covariates to adjust for potential confounders.

All statistical analyses were performed using Stata software (Stata Statistical Software: Release 16; StataCorp LLC, TX, USA). A p-value of <0.05 was considered statistically significant.

## Results

Table 1 presents the characteristics of the participants. The prevalence rate of LBP in the study participants was 49.9%. Compared to the participants without LBP, those who experienced LBP were younger, middle-aged workers (20–49 years: 43.1% vs. 52.2%), and predominantly females (37.6% vs. 48.2%).

**Table 1.**
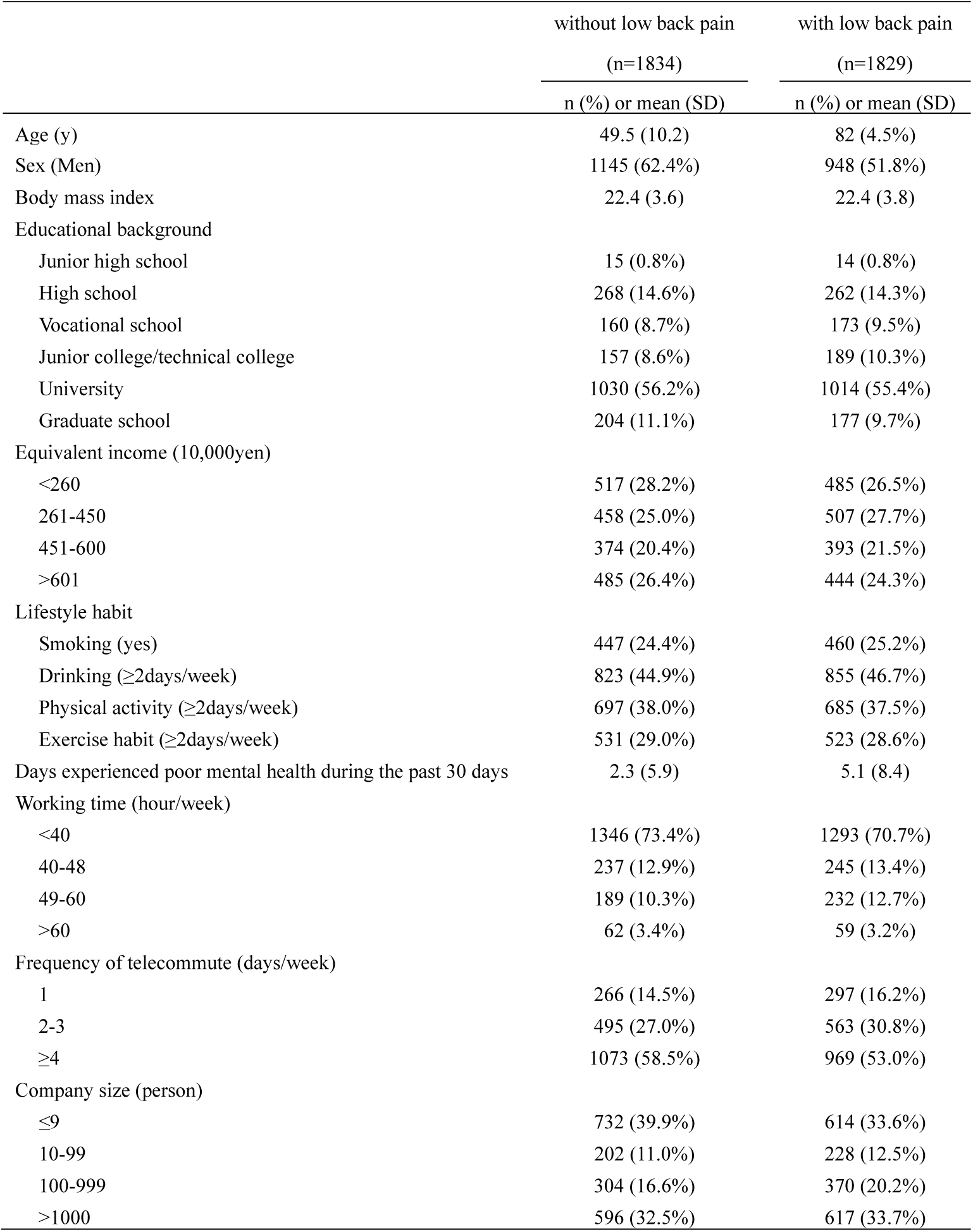
Characteristics of participants

Table 2 shows the telecommuting environment of the participants. The proportion of participants who answered “no” to each question regarding their telecommuting environment was higher among participants with LBP than among those without LBP.

**Table 2.** Telecommuting environment of participants

|  | without low back pain<br>(n=1834) | with low back pain<br>(n=1829) |
| --- | --- | --- |
|  | n (%) | n (%) |
| Telecommuting environment |  |  |
| Is your desk well-lit for you to work? (No) | 235 (12.8%) | 361 (19.7%) |
| Is there enough space to stretch the legs? (No) | 334 (18.2%) | 450 (24.6%) |
| Do you have a place or room where you can concentrate on your work? (No) | 351 (19.1%) | 496 (27.1%) |
| Are the degrees of temperature and humidity in the room where you work appropriate for working comfortably? (No) | 395 (21.5%) | 526 (28.8%) |
| Do you have enough space on your desk to work? (No) | 472 (25.7%) | 582 (31.8%) |
| Do you use an office desk or chair? (Including children's study desks) (No) | 860 (46.9%) | 931 (50.9%) |

The association between LBP and the telecommuting environment is shown in Table 3. There was a significant association between LBP and the following questions: “Is your desk well-lit enough for you to work? (No)” (odds ratio [OR]: 1.43, 95% confidence interval [CI]: 1.18–1.73, p<0.001), “Is there enough space to stretch the legs? (No)” (OR: 1.30, 95% CI: 1.10–1.54, p=0.003), “Do you have a place or room where you can concentrate on your work? (No)” (OR: 1.38, 95% CI: 1.17–1.63, p<0.001), “Are the degrees of temperature and humidity in the room where you work appropriate for working comfortably? (No)” (OR: 1.32, 95% CI: 1.13–1.56, p=0.001), and “Do you have enough space on your desk to work? (No)” (OR: 1.19, 95% CI: 1.02–1.39, p=0.024). There was no significant association between LBP and the question “Do you use an office desk or chair? (No)” (OR: 1.03, 95% CI: 0.90–1.18, p=0.696). Similar results were obtained for Model 1, which was adjusted only for sex and age, and for Model 2, which was adjusted for other potential confounders.

**Table 3.** The association between low back pain and telecommuting environment

|  | Model 1<sup>†</sup> |  |  |  | Model 2<sup>‡</sup> |  |  |  |
| --- | --- | --- | --- | --- | --- | --- | --- | --- |
|  | OR | 95% CI |  | P | OR | 95% CI |  | P |
| Telecommuting environment |  |  |  |  |  |  |  |  |
| Is your desk well-lit **enough** for you to work? (No) | 1.66 | 1.38 | 1.99 | <0.001 | 1.43 | 1.18 | 1.73 | <0.001 |
| Is there enough space to stretch the legs? (No) | 1.47 | 1.25 | 1.73 | <0.001 | 1.30 | 1.10 | 1.54 | 0.003 |
| Do you have a place or room where you can concentrate on your work? (No) | 1.53 | 1.31 | 1.80 | <0.001 | 1.38 | 1.17 | 1.64 | <0.001 |
| Are the degrees of temperature and humidity in the room where you work appropriate for working comfortably? (No) | 1.45 | 1.25 | 1.69 | <0.001 | 1.32 | 1.13 | 1.56 | 0.001 |
| Do you have enough space on your desk to work? (No) | 1.34 | 1.16 | 1.55 | <0.001 | 1.19 | 1.02 | 1.39 | 0.024 |
| Do you use an office desk or chair? (Including children's study desks) (No) | 1.11 | 0.97 | 1.27 | 0.121 | 1.03 | 0.90 | 1.18 | 0.696 |

<sup>†</sup>Model 1 adjusted for age and sex
<sup>‡</sup>Model 2 adjusted for age, sex, body mass index, lifestyle habit (smoking, drinking, physical activity, and exercise habit), number of days the participants experienced poor mental health during the past 30 days, equivalent income, educational background, working time, frequency of telecommuting, and company size

## Discussion

This study showed that telecommuting environment was associated with LBP in home workers during the COVID-19 pandemic. Particularly, it was suggested that insufficient desk and foot space, inadequate desk lighting, uncomfortable room temperature and humidity, and lack of room/space for concentrating on work were associated with the prevalence of LBP.

In this study, workspace-related factors (e.g., room brightness, temperature, and humidity) had a stronger impact on LBP than the environment around the desk being used (e.g., desk, chair, foot space). Because brightness and temperature/humidity are properly controlled in a typical office, attention tends to be focused on the environment around the desk (e.g., desk, chair, etc.), when considering LBP and the work environment of desk workers. However, in the case of telecommuting, control of brightness, temperature, and humidity is left to the worker and may not be properly managed. The results of this study suggest the importance of paying attention to the entire workspace when considering LBP prevention among telecommuters.

In this study, inadequate desk and foot space, and insufficient lighting were associated with LBP. It is suggested that an awkward posture and sitting for long durations are risk factors for LBP^8,22–24^. Having enough space on the desk and for the feet is effective in maintaining good posture when working as well as changing the posture and stretching as needed. In addition, inadequate lighting at the desk may contribute to awkward postures among workers when looking at documents or computer screens on their desk. It is necessary to manage the work environment to avoid awkward postures and prolonged maintenance of the same posture during work.

The present study also revealed that uncomfortable temperature and humidity in the telecommuting space was associated with LBP. Because this study was conducted during the winter season, it can be speculated that the results imply that cold temperature and low humidity are associated with LBP. The relationship between cold temperature and LBP has been shown in previous studies^15,25–27^, which is the same for telecommuting workers. Although the most suitable room temperature for the prevention of LBP is not clear, we consider that room temperatures that are subjectively cold should be avoided, as they may increase the risk of musculoskeletal symptoms and injuries.

In this study, using an office chair/desk was not associated with LBP. This finding is supported by those of previous studies reporting no significant association between LBP and use of chair/desk in office workers^11,28^. Alternatively, previous studies have reported that LBP is related to the characteristics of the chair, such as with or without lumbar support and adjustable back support^8,22^. This may suggest that it is not only a matter of whether an office chair is used but also what function and shape of the chair is used for the prevention of LBP.

The lack of room or space to concentrate on work was associated with LBP. It is suggested that the teleworker’s workstation should be in a dedicated space that is private, quiet, and secure, preferably away from the flow of activity in the home^4^. The results of this study support that argument. Previous studies have reported that psychological stress is associated with LBP in workers^29–31^. Lack of a room or space to concentrate on work may cause psychological stress to telecommuting workers. The results of this study suggest that it is important for telecommuting workers to have a space where they can devote themselves to their work as much as possible, despite working from home.

Based on these findings, we suggest that the work environment of telecommuting workers may be associated with LBP. Therefore, employers should educate telecommuting workers about the importance of an appropriate home working environment. If it is difficult to prepare an appropriate telecommuting environment due to household situations and family structure, then it is necessary to consider the use of co-working spaces, satellite offices, and spaces near the employee’s residence for telecommuting.

This study has three limitations. First, specific details such as history, symptom duration, and LBP diagnosis are unknown, as only the NRS was used for evaluation. However, it is uncertain how these factors affect the relationship between the telecommuting environment and LBP. Second, home environment, lifestyle factors, and working condition were also evaluated using subjective questions; hence, the validity of the responses is unclear. However, an objective method to evaluate the work environment at home has not been established at this time. Third, selection bias may have occurred in this study. People with LBP may be more likely to choose telecommuting if possible. However, the effect of this bias on the results of this study is uncertain.

## Conclusion

The present study suggests that telecommuting environment is associated with the prevalence of LBP among telecommuting workers in Japan, indicating that employers may need to educate their employees to maintain a healthy and ergonomic environment.

## Data Availability

Data not available due to ethical restrictions

## Acknowledgements

This study was supported and partly funded by the research grant from the University of Occupational and Environmental Health, Japan (no grant number); Japanese Ministry of Health, Labour and Welfare (H30-josei-ippan-002, H30-roudou-ippan-007, 19JA1004, 20JA1006, 210301-1, and 20HB1004); Anshin Zaidan (no grant number), the Collabo-Health Study Group (no grant number), and Hitachi Systems, Ltd. (no grant number) and scholarship donations from Chugai Pharmaceutical Co., Ltd. (no grant number)

The current members of the CORoNaWork Project, in alphabetical order, are as follows: Dr. Yoshihisa Fujino (present chairperson of the study group), Dr. Akira Ogami, Dr. Arisa Harada, Dr. Ayako Hino, Dr. Hajime Ando, Dr. Hisashi Eguchi, Dr. Kazunori Ikegami, Dr. Kei Tokutsu, Dr. Keiji Muramatsu, Dr. Koji Mori, Dr. Kosuke Mafune, Dr. Kyoko Kitagawa, Dr. Masako Nagata, Dr. Mayumi Tsuji, Ms. Ning Liu, Dr. Rie Tanaka, Dr. Ryutaro Matsugaki, Dr. Seiichiro Tateishi, Dr. Shinya Matsuda, Dr. Tomohiro Ishimaru, and Dr. Tomohisa Nagata. All members are affiliated with the University of Occupational and Environmental Health, Japan.

